# Personality Profiles in Bipolar Disorder: Differences Across Diagnostic Subtypes and Associations with Demographic and Health Factors

**DOI:** 10.64898/2026.08.06.26359885

**Authors:** Lucía Albarracín-García, Inés García-Ortiz, Alejandro Porras-Segovia, Laura Navío-García, Laura Jiménez-Muñoz, Elisabet Madridejos-Palomares, Beatriz M. Gonzalez-Toledo, Olatz López-Fernández, Enrique Baca-García, Claudio Toma

**Author notes:** Corresponding authors: Alejandro Porras-Segovia, University Hospital Jimenez Diaz Foundation, Madrid, Spain,; Claudio Toma, Centro de Biología Molecular (CBMSO), Calle Nicolás Cabrera 1, 28049 Madrid, Spain. equally contributing first authors (Lucía Albarracín-García, Inés García-Ortiz). equally contributing senior authors (Enrique Baca-García, Claudio Toma).

## Abstract

**Background:** Personality traits are consistently associated with bipolar disorder (BD). However, their features across BD diagnostic subtypes and their modulation by demographic and health-related factors remain largely unexplored. This study aimed to characterize Big Five personality domains in individuals with BD compared to controls, and to examine differences between BD type I (BD-I) and BD type II (BD-II).

**Methods:** We analyzed 833 participants from the MadManic cohort (300 BD subjects and 533 controls) with available Big Five Inventory-2 (BFI-2) data. Linear regression models were used to assess associations between personality traits and BD diagnosis, adjusting for relevant covariates. Additional comparisons were conducted across sex, age, and Body Mass Index (BMI), and between BD-I and BD-II patients.

**Results:** BD was associated with higher *Negative Emotionality* (*NE*) and lower *Extraversion* and *Conscientiousness*. *Conscientiousness* was also inversely associated with BMI. Within the BD group, individuals with BD-I exhibited lower *NE* compared to those with BD-II. Stratified analyses indicated that elevated *NE* in BD was the most consistent domain across sex, age, and BMI subgroups, whereas differences in *Extraversion* and *Conscientiousness* varied depending on subgroup features.

**Conclusions:** BD is characterized by a distinct personality profile marked by elevated *NE* and reduced *Extraversion* and *Conscientiousness. NE* emerged as the most robust domain associated with BD, which may also differentiate between subtypes, with higher levels observed in BD-II than BD-I. These findings highlight the relevance for considering demographic and health-related factors, particularly BMI, when interpreting personality patterns in BD, supporting the role of personality dimensions to examine clinical heterogeneity.

## INTRODUCTION

Bipolar Disorder (BD) is a severe and chronic psychiatric condition characterized by recurrent episodes of mania, hypomania, and depression, and is associated with substantial functional impairment and reduced quality of life (Grande et al., 2016). Beyond its episodic nature, BD exhibits marked clinical heterogeneity in terms of age at onset, episode polarity, comorbidities, illness course, and treatment response (Vieta et al., 2018). Importantly, many individuals with BD experience residual symptoms and functional difficulties even during euthymic periods, suggesting the presence of enduring trait-like vulnerabilities (Samalin et al., 2016). Despite advances in diagnosis and treatment, BD remains a leading contributor to global disability, underscoring the need to identify factors that account for this variability (Yocum et al., 2026). In this context, personality may represent a relevant dimension for understanding the heterogeneity and clinical expression of BD.

Dimensional models of personality provide a robust framework for capturing individual differences relevant to psychiatric vulnerability (McAdams, 1992; Trull and Widiger, 2013). The Five-Factor Model (FFM), commonly operationalized using instruments such as the Big Five Inventory-2 (BFI-2), characterizes personality across five broad domains: *Extraversion*, *Openness to experience*, *Conscientiousness*, *Agreeableness* and *Neuroticism*, referred to as *Negative emotionality* (NE) in the BFI-2 (Soto and John, 2017). These traits are continuously distributed in the general population and reflect relatively stable patterns of emotional and behavioural functioning across adulthood, in which sex-differences, behaviours, and health factors, such as body mass index (BMI), play a key role as modulators (Sutin et al., 2011; Bogg and Roberts, 2004; Costa et al., 2001). Indeed, several evidence indicate that age and personality are mutually influenced by each other: *Conscientiousness* and *Agreeableness* increase over time, while *Neuroticism* decreases. In addition, behavioural and personality patterns are shaped not only by developmental changes but also by a generational imprinting (Brandt et al., 2022).

The association of FFM dimensions with behavioural profiles makes them particularly relevant for understanding disease risk, clinical heterogeneity, and course modifiers in psychiatric disorders (Cobb-Clark and Schurer, 2012).

The study performed by Furukawa *et al*. (1998), was one of the first research to assess aspects of personality traits in psychiatric disorders using the FFM. The findings indicated that overall psychiatric patients showed higher levels of *Neuroticism* and lower levels of *Conscientiousness* compared to controls. No specific personality traits were found to be clearly associated to a psychiatric diagnosis, although the sample size was relatively small (N=140), preventing from general conclusions (Furukawa et al., 1998). Subsequent research has shifted from the broad psychiatric category to more targeted analyses into individual diagnosis. For instance, some authors have explored personality traits in schizophrenia (Ohi et al., 2016), while others have moved toward analysing psychiatric symptoms (Malouff et al., 2005) or trans-diagnostic factors, such as the internalizing dimension shared by anxiety and depression (Griffith et al., 2010, Kotov et al., 2010).

Accumulating evidence suggests that personality traits and psychiatric disorders may share genetic and biological underpinnings (Smeland et al. 2017). Genetic studies indicate partial overlap between the genetic architecture of personality traits, and *Neuroticism* in particular, with psychiatric phenotypes (Grotzinger et al., 2022; Streit et al., 2022).

However, this overlap appears to be partial, indicating that personality traits capture only some aspects of the underlying biological vulnerability. Indeed, other studies have not found a genetic association between personality traits and specific clinical outcomes, such as suicidal behaviour (Kalman et al., 2022).

Within BD, several studies have identified distinct personality profiles compared to general population. Results consistently indicates that BD is associated with specific trait patterns. Hanke et al. conducted a meta-analysis, including 18 studies (1,694 BD patients and 2,153 controls) and found that individuals with BD exhibit higher levels of *Neuroticism* and lower levels of *Extraversion* and *Conscientiousness* (Hanke et al., 2022).

Similar personality patterns were reported across other additional studies, including cross-diagnostic (Bagby et al., 1997) and longitudinal mood assessment studies (Barnett et al., 2011; Sparding et al., 2017; Ryan et al., 2021; Fleischmann et al., 2023). Evidence suggest that personality domains show relative stability over time in BD populations. Indeed, Ryan et al. found that elevated *Neuroticism*, reduced *Extraversion* and *Conscientiousness* persist across follow-up interviews, although *Neuroticism* appears to be more sensitive to fluctuations during depressive symptoms (Ryan et al. 2025; Ryan et al. 2021).

Comparisons across psychiatric phenotypes further support the trans-diagnostic relevance of personality trait measures. While high *Neuroticism* and low *Extraversion* are frequently cited as core markers of BD (Jylhä et al., 2010; Quilty et al., 2009; Almeida et al., 2011), other domains like *Openness* and *Agreeableness* show greater heterogeneity across studies. Other studies aimed to identify specific personality patterns able to distinguish across diagnostic groups. In mood disorders, some studies identified differences between BD and depression based on *Extraversion*, *Conscientiousness* or *Neuroticism* levels (Canuto et al., 2010; Hirschfeld et al., 1979; Li et al., 2024), while other studies found that both conditions exhibit basically similar personality elevations (Jylhä et al., 2010; Li et al., 2024).

In psychotic disorders a recent meta-analysis suggests that schizophrenia is characterized by more pronounced alterations in lower *Extraversion*, *Openness*, and *Agreeableness* compared to BD (Hashimoto et al., 2025), highlighting that both have distinct personality profiles, despite sharing symptoms and genetic susceptibility. Komasi *et al*. found that BD patients were different from borderline personality disorder (BPD) patients regarding *Neuroticism*, *Extraversion*, *Openness*, and *Agreeableness*, while no difference was observed for *Conscientiousness* (Komasi et al., 2022). This meta-analysis showed also that BD and BPD were different when compared with general population in all FFM factors, except for *Openness* between BD and controls (Komasi et al., 2022). Other studies focused on personality functioning between BPD and BD describing greater impairment in BPD, especially in personality organization (Feichtinger et al., 2024).

While these mentioned studies focused mainly on the comparisons between BD and controls, or BD and other major psychiatric conditions, only few studies have explored potential differences in personality traits in BD subtypes. A study by Kim *et al*., suggested higher *Neuroticism* and lower *Extraversion* in BD-II (N=43) compared to BD-I (N=85) (Kim et al., 2012), although the sample size was small. Another study found that BD-I shows higher *Openness* than BD-II in a Chinese version of the BFI in a sample of 252 BD patients (Lin et al., 2025).

Despite the growing interest in this area, several aspects of personality traits need to be better explored in BD: i) The adoption of more improved instruments such as the Big Five Inventory-2 (BFI-2) in large and well-characterized BD cohorts; ii) Comparison between BD-I and BD-II subtypes that may highlight specific patterns otherwise undetected; iii) Take into account clinically relevant variables for clinical course, such as sex, age, and the body mass index (BMI) as general indicator of health conditions.

The present study aims to address these gaps by examining personality traits, as defined by the BFI-2, in a large sample of BD cases, clinically assessed and compared with a control population. Additionally, we explore differences between BD subtypes and assess the effect of demographic and clinical variables, including sex, age, and BMI.

## MATERIALS AND METHODS

### Cohort and BFI-2 administration

This study comprises BD cases and controls from the *MadManic* cohort (García-Ortiz *et al*., 2026), a sample of BD patients and controls from Spain for whom extensive biological, clinical and behavioural data have been collected. Approvals to handle clinical or behavioural scales of BD patients and controls were obtained by the research ethics committee of the Spanish National Research Council (Consejo Superior de Investigaciones Científicas, CSIC) (13/2021; 109/2023; 030/2025) and Fundación Jiménez Díaz Hospital (15/21; 11/23) (García-Ortiz *et al*., 2026). All participants provided written informed consent.

The BFI-2, a 60-item personality inventory assessing five broad personality domains, as well as three narrower facets within each domain (Soto and John, 2017), was administered to BD patients and controls. A random subset of controls was selected to match the age distribution of the BD group in order to minimize the effect of generational variability in personality expression. All the BD participants were in a euthymic state at the time of participation. Sociodemographic and BMI data were collected through face-to-face interviews or via self-administered written questionnaires.

### Statistical analyses

All statistical analyses were conducted using R (v.4.3.0) and the psych R package (v.2.4.2). Statistical significance was set at P-value < 0.05. Bonferroni-corrected P-value was specified when required to account for multiple testing.

Demographic characteristics were summarized, and group comparisons between BD cases and controls, as well as between BD subtypes, were performed using the Wilcoxon rank-sum test for continuous variables and the χ² test for categorical variables. Continuous variables showing significant group differences were visually examined using kernel density plots. Relationships between covariates (sex, age, BMI, ethnicity, BD diagnosis and BD subtype) and BFI-2 domain scores were calculated using the Pearson correlation. The overall scale, as well as the personality domains, was evaluated using Cronbach’s alpha.

Unadjusted comparisons between BD and control groups, as well as between BD-I and BD-II, were conducted for the five BFI-2 domains. Effect sizes were quantified using Cohen’s *d*, and group differences were evaluated using the Wilcoxon rank-sum test. The contribution of individual facets was assessed using the same statistical approach.

To assess the association between BD status and BFI-2 domain scores while accounting for potential confounders, multivariable linear regression models were fitted separately for each domain. Each model included sex (male or female), age, BMI, and diagnostic group (control or BD). Potential interactions between diagnostic group and sex, age, or BMI were initially evaluated; as none improved model fit or reached statistical significance, final models retained only additive effects. BFI-2 domain scores were also compared between BD cases and controls within predefined subgroups, according to sex (male or female), age (<40 or ≥40 years) and BMI (<25 or ≥25 kg/m², the conventional threshold for overweight). Effect sizes (Cohen’s *d*) and Wilcoxon P-values are reported.

## RESULTS

### Sociodemographic characteristics

A total of 833 participants were included in the study, comprising 300 BD cases and 533 controls (**Table 1**). Amongst the BD participants, 161 were diagnosed with BD-I, 95 with BD-II, 23 with schizoaffective disorder bipolar type (SZMA), and 21 under a BD spectrum (BS) (**Supplementary Table 1**). Age and BMI differed significantly between BD cases and controls (*P* = 5.27E-07 and *P* = 3.36E-16, respectively), with BD participants showing higher mean age (54.60 vs. 50.46 years) and BMI (27.99 vs. 24.97). Kernel density plots further illustrated a shift toward higher values for both variables in the BD group (**Fig. 1**), consistent with reports of higher prevalence of overweight and obesity in BD individuals (Vancampfort et al., 2015). Despite these differences, the proportion of individuals younger than 40 years was comparable between groups (**Fig. 1A**). This threshold was used descriptively as a lower-liability class for BD onset, since control participants above this age are less likely to subsequently develop the disorder (Toma et al., 2018). Correlation analyses showed weak association between age and BMI (Pearson’s r = 0.14; **Supplementary Fig. 1**).

**Figure 1:**
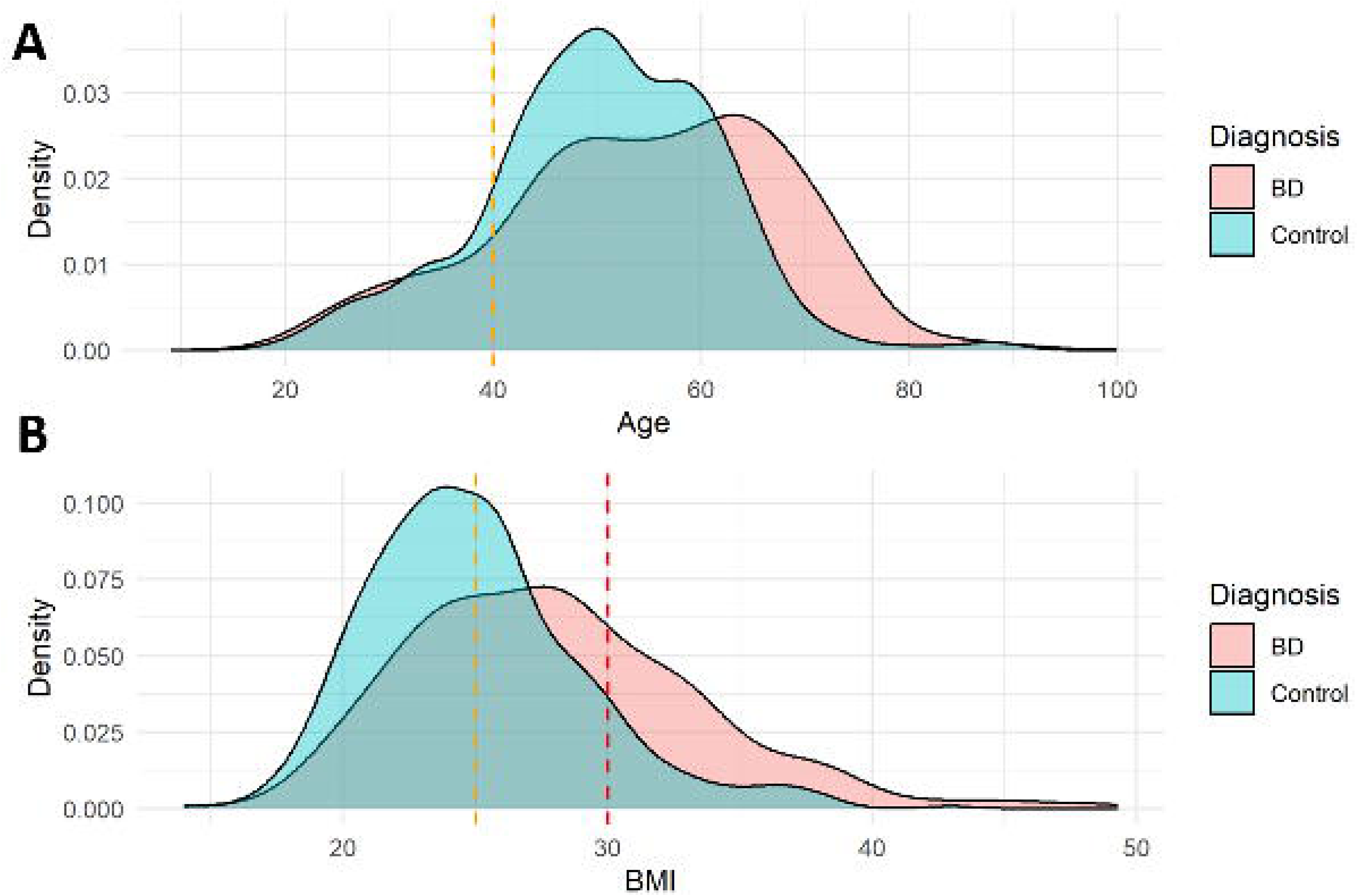
Kernel density distributions of age and BMI by diagnostic group. **A)** Age distribution in BD cases and controls. Dashed orange line indicates the 40 years descriptive threshold for lower BD-onset liability in controls; **B)** BMI distribution in BD cases and controls. Orange dashed line at BMI = 25 kg/m², indicating the threshold for overweight, and red dashed line at BMI = 30 kg/m², indicating the threshold for obesity.

**Table 1:** General characteristics of the study sample. Continuous variables are reported as mean ± standard deviation and were compared between BD cases and controls using Wilcoxon rank-sum tests. Categorical variables are reported as counts and percentages and were compared using chi-squared tests. BD subtypes are reported within the BD group only. P-values shown in bold indicate statistically significant results after Bonferroni correction, using a corrected significance threshold of P < 0.01.

|  | Total (N = 833) |  | BD (N = 300) |  | Controls (N=533) |  | Statistic | P |
| --- | --- | --- | --- | --- | --- | --- | --- | --- |
| | N (%) | Mean ± SD | N (%) | Mean ± SD | N (%) | Mean ± SD | | |
| Females | 477 (57.3) | | 188 (62.7) | | 289 (54.2) | | $X^2 = 5.25$ | 0.022 |
| Age | | 51.95 ± 13.21 | | 54.60 ± 13.68 | | 50.46 ± 10.98 | W = 63233 | **5.27E-07** |
| Age group <40 years | 130 (15.6) | | 47 (15.6) | | 83 (15.6) | | $X^2 = 2E-29$ | 1 |
| BMI | | 26.06 ± 4.85 | | 27.99 ± 5.48 | | 24.97 ± 4.09 | W = 52749 | **3.36E-16** |
| Non-European | 47 (5.6) | | 20 (6.7) | | 27 (5.1) | | $X^2 = 0.64$ | 0.421 |
| BD-I |  |  | 161 (53.7) |  |  |  |  |  |
| BD-II |  |  | 95 (31.7) |  |  |  |  |  |
| SZMA |  |  | 23 (7.7) |  |  |  |  |  |
| BS |  |  | 21 (6.9) |  |  |  |  |  |

Abbreviations: BD-I, bipolar disorder type I; BD-II, bipolar disorder type II; SZMA, schizoaffective disorder bipolar type; BS, Bipolar Spectrum; BMI, Body Mass Index; SD, Standard Deviation; W, Wilcoxon statistic; $\chi^2$ , chi-squared statistic; P, P-value

### Personality domains

The overall internal consistency for the entire BFI-2 scale was high (Cronbach’s α = 0.90), as well as for all five domains (Cronbach’s α >0.80) (**Supplementary Table 2**), suggesting excellent internal reliability of the scale in our study. Correlation analyses showed that associations between BFI-2 domain scores and covariates were generally small (**Supplementary Fig. 2**). In this regard, *Negative Emotionality* (*NE*) was positively correlated with BD diagnosis (Pearson’s r = 0.14), with weaker positive associations with age (Pearson’s r = 0.06) and BMI (Pearson’s r = 0.08). Conversely, *Conscientiousness* showed the largest correlation being negatively associated with BMI (Pearson’s r = -0.18) (**Supplementary Fig. 2**).

Comparing personality domains, individuals with BD showed significantly higher scores in *NE* compared to controls (*P*<2.2E-16), and lower scores in *Extraversion* (*P*=1.9E-10) and *Conscientiousness* (*P*=8.12E-07) (**Table 2**). Facet-level analysis indicated that the domain differences were largely consistent across their underlying facets (**Supplementary Table 3**). Within *Extraversion*, BD participants scored lower than controls in *Sociability*, *Assertiveness*, and *Energy Level*. Within *Conscientiousness*, lower scores were observed in *Productivity* and *Responsibility*, whereas *Organization* did not differ between groups. Conversely, all *NE* facets, including *Anxiety*, *Depression*, and *Emotional Volatility*, were higher in BD participants than in controls. A significant group difference was also observed for the *Intellectual Curiosity* facet, with lower scores in individuals with BD (*P*=2.79E-06), despite the absence of a significant differences in the overall *Openness* domain.

**Table 2:** Unadjusted comparisons of BFI-2 domain scores between BD patients and controls. Values are shown as mean ± standard deviation. Effect sizes are reported as Cohen’s d, and P-values correspond to Wilcoxon rank-sum tests. P-values surviving Bonferroni correction are shown in bold.

| Trait | Total<br>(N=833) | BD<br>(N=300) | Controls<br>(N=533) | Cohen's d | P |
| --- | --- | --- | --- | --- | --- |
| *Extraversion* | 3.39 ± 0.69 | 3.20 ± 0.74 | 3.52 ± 0.61 | 0.490 | **1.9E-10** |
| *Agreeableness* | 4.13 ± 0.51 | 4.11 ± 0.57 | 4.15 ± 0.47 | 0.089 | 0.465 |
| *Conscientiousness* | 3.68 ± 0.70 | 3.49 ± 0.78 | 3.77 ± 0.62 | 0.402 | **8.12E-07** |
| *Negative*<br>*Emotionality* | 2.87 ± 0.78 | 3.25 ± 0.82 | 2.65 ± 0.67 | -0.837 | **<2.2E-16** |
| *Openness* | 3.80 ± 0.69 | 3.72 ± 0.78 | 3.84 ± 0.63 | 0.177 | 0.074 |

Abbreviations: BD, Bipolar Disorder; BFI-2, Big Five Inventory-2; SD, Standard Deviation; P, P-value.

A BD subtype analysis was conducted in the subset of individuals with BD-I or BD-II diagnoses (N = 256). In these comparisons, BFI-2 domain scores were broadly similar between subtypes (**Table 3**). The only nominal difference was observed for *NE*, with BD-I patients scoring lower than BD-II patients (*P* = 0.014).

**Table 3:**
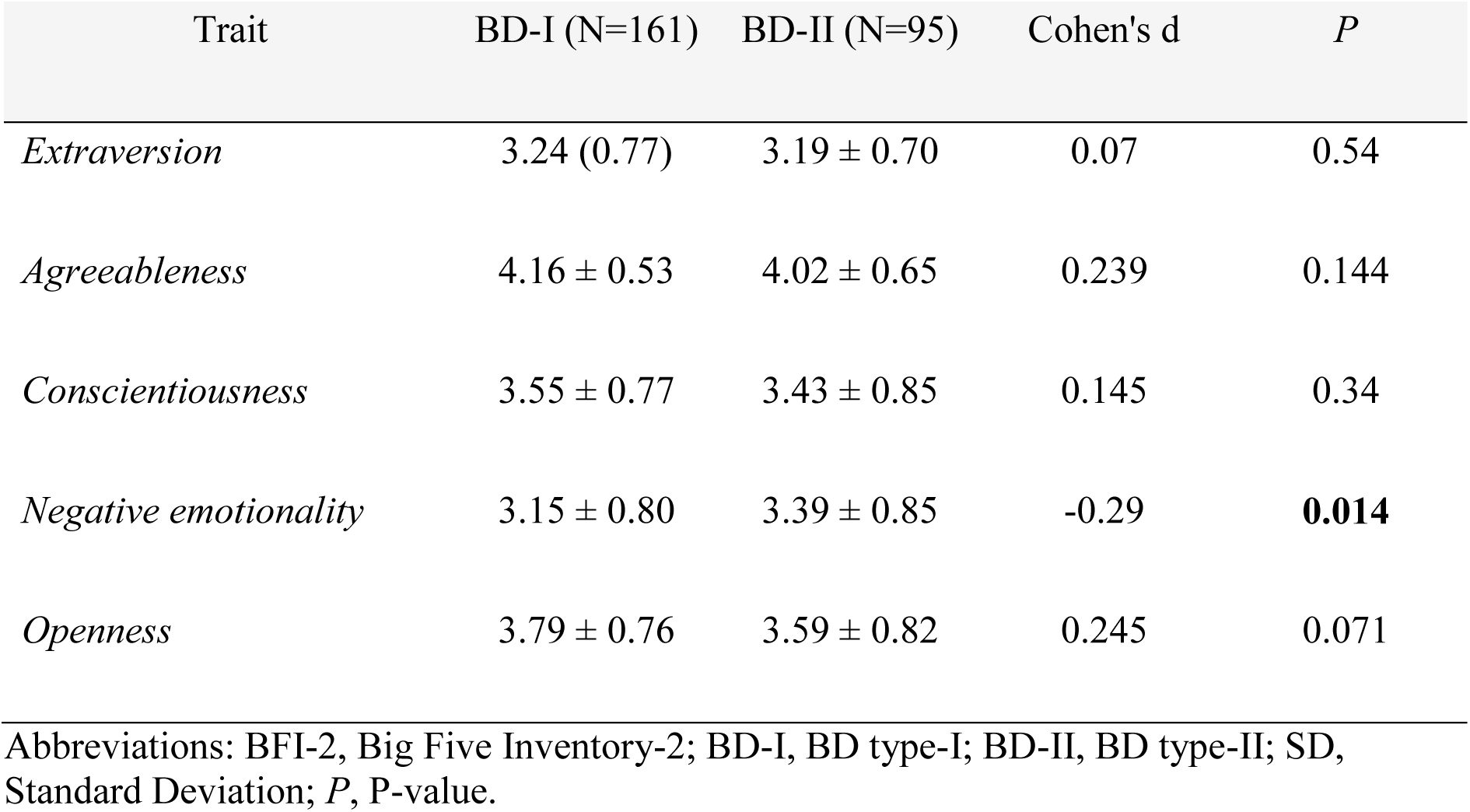
Unadjusted comparisons of BFI-2 mean scores between BD-I and BD-II patients. Values are shown as mean ± standard deviation. Effect sizes are reported as Cohen’s d, and P-values correspond to Wilcoxon rank-sum tests.

| Trait | BD-I (N=161) | BD-II (N=95) | Cohen's d | P |
| --- | --- | --- | --- | --- |
| *Extraversion* | 3.24 (0.77) | 3.19 ± 0.70 | 0.07 | 0.54 |
| *Agreeableness* | 4.16 ± 0.53 | 4.02 ± 0.65 | 0.239 | 0.144 |
| *Conscientiousness* | 3.55 ± 0.77 | 3.43 ± 0.85 | 0.145 | 0.34 |
| *Negative emotionality* | 3.15 ± 0.80 | 3.39 ± 0.85 | -0.29 | **0.014** |
| *Openness* | 3.79 ± 0.76 | 3.59 ± 0.82 | 0.245 | 0.071 |

Abbreviations: BFI-2, Big Five Inventory-2; BD-I, BD type-I; BD-II, BD type-II; SD, Standard Deviation; P, P-value.

### Personality domains differences in BD and modulation by sex, age and BMI

Additive multiple linear regression models were fitted separately for each personality domain to account for the effects of BD diagnosis, sex, age, and BMI (**Table 4**). BD diagnosis remained associated with lower *Extraversion* and *Conscientiousness* (*P*=5.91E-11 and *P*=5.77E-07, respectively), and higher *NE* (*P*<2E-16). No significant association with BD diagnosis was observed for *Agreeableness* or *Openness*. Beyond the effect of BD diagnosis, each personality domain showed a distinct pattern of association with these other traits. *Extraversion* was only weakly related to sex and age at a nominal level. *NE* was mainly associated with diagnosis, but considerably also with sex, whereas age and BMI showed no contribution to this domain. *Conscientiousness* showed the broadest covariate modulation, with significant associations with sex, age, and BMI, a part with diagnosis, suggesting that this domain is particularly modulated by demographic and health risk factors. In contrast, *Openness* was mainly related to age, showing lower scores with increasing age, and *Agreeableness* to sex, showing higher scores in females.

**Table 4:** Multivariable linear regression models assessing the association between BD diagnosis and BFI-2 personality domain scores, adjusted for sex, age, and BMI. Separate models were fitted for each personality domain. β estimates represent the adjusted effect of each predictor on the corresponding BFI-2 domain score. Statistical significance after Bonferroni correction was set at P < 0.0025; values meeting this threshold are shown in bold.

| Trait | Predictor | Estimate ($\beta$) | S.E. | t | P |
| --- | --- | --- | --- | --- | --- |
| *Extraversion* | Sex | 0.137 | 0.047 | 2.91 | 0.004 |
|  | Age | -0.004 | 0.002 | -2.28 | 0.023 |
|  | BMI | 0.006 | 0.005 | 1.16 | 0.246 |
|  | Diagnosis | -0.334 | 0.050 | -6.63 | **5.91E-11** |
| *Agreeableness* | Sex | 0.169 | 0.036 | 4.71 | **2.95E-06** |
|  | Age | -0.002 | 0.001 | -1.66 | 0.097 |
|  | BMI | -0.002 | 0.004 | -0.53 | 0.599 |
|  | Diagnosis | -0.043 | 0.038 | -1.13 | 0.260 |
| *Conscientiousness* | Sex | 0.163 | 0.048 | 3.39 | **7.34E-04** |
|  | Age | 0.006 | 0.002 | 3.25 | **0.001** |
|  | BMI | -0.019 | 0.005 | -3.64 | **2.91E-04** |
|  | Diagnosis | -0.258 | 0.051 | -5.04 | **5.77E-07** |
| *Negative emotionality* | Sex | 0.232 | 0.052 | 4.47 | **8.78E-06** |
|  | Age | -0.001 | 0.002 | -0.67 | 0.505 |
|  | BMI | -0.002 | 0.006 | -0.32 | 0.748 |
|  | Diagnosis | 0.601 | 0.055 | 10.87 | **< 2E-16** |
| *Openness* | Sex | 0.066 | 0.049 | 1.35 | 0.179 |
|  | Age | -0.006 | 0.002 | -3.18 | **1.53E-03** |
|  | BMI | -0.002 | 0.005 | -0.37 | 0.714 |
|  | Diagnosis | -0.095 | 0.052 | -1.81 | 0.071 |

Abbreviations: BFI-2, Big Five Inventory-2; BD, Bipolar Disorder; BMI, Body Mass Index; S.E., Standard Error; t, t-value; P, P-value.

To further characterize differences in personality patterns beyond affection status, subgroup analyses were conducted for sex, age, and BMI comparing BD subjects to controls (**Fig. 2**, **Supplementary Table 4**). The threshold for significance was set to *P* < 0.0017, after considering Bonferroni correction. In sex-stratified analyses, both male and female BD participants showed lower *Extraversion* and higher *NE* than controls, whereas lower *Conscientiousness* survived Bonferroni correction only among females (**Fig. 2A**). When stratifying the sample by age using a 40-year threshold, higher *NE* was observed in BD cases in both age groups, while lower *Extraversion* and *Conscientiousness* were restricted to participants older than 40 years (**Fig. 2B**). In BMI-stratified analyses, higher *NE* was observed in BD cases across both BMI categories, whereas lower *Extraversion* and *Conscientiousness* survived correction only among participants with BMI >25 kg/m² (**Fig. 2C**). No significant differences were observed for *Agreeableness* or *Openness* in any subgroup. However, nominal differences in *Openness* were observed in the age and BMI stratified analyses. Interestingly, higher scores of *Openness* were observed in BD cases compared to controls in the younger group; while this differences remained significant in the older group, in this case BD subjects showed an opposite direction with lower scores compared to controls.

**Figure 2:**
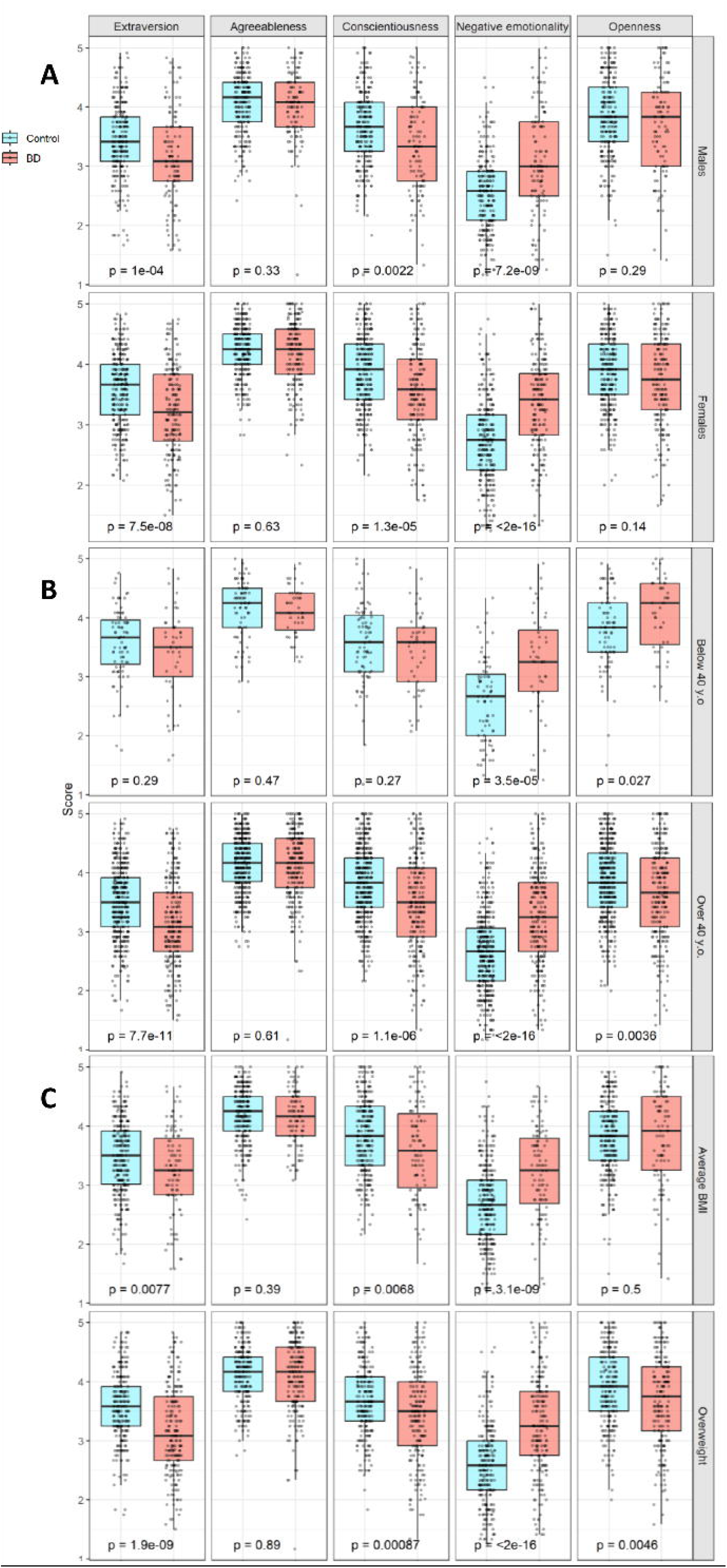
**Boxplots of BFI-2 domain scores in BD cases and controls across predefined subgroups**. Comparison are shown by: A) Sex (males, N = 356; females, N = 477); B) Age category (≤40 years, N = 130; >40 years, N = 703); C) BMI category (BMI ≤25 kg/m², N = 407; BMI >25 kg/m², N = 426). Individual data points are overlaid, and P-values indicate Wilcoxon rank-sum tests comparing BD cases and controls within each subgroup and personality domain.

Overall, these stratified analyses indicate that increased *NE* was the most consistent feature of BD across sex, age, and BMI subgroups, whereas differences in *Extraversion* and *Conscientiousness* were more dependent on subgroup structure.

## DISCUSSION

The characterization of personality traits can help to clarify the substantial clinical heterogeneity observed in BD by identifying endophenotypes related to biological dysregulated mechanisms, distinguishing between clinical subgroups, and predicting clinical course and outcomes. Previous studies reported significant differences in Big Five personality domains between BD and general population, as well as BD and other psychiatric conditions. However, many of these studies were conducted in relatively small cohorts, including approximately 200 BD patients or fewer (Li et al., 2024; Sparding et al., 2017; Kim et al., 2012; Canuto et al., 2010; Furukawa et al., 1998; Bagby et al., 1997). To date, there are no studies that have systematically examined whether personality patterns in BD vary across demographic and health-related categories. In the present study, we extend previous work by examining BFI-2 personality domains and facets both in the overall BD diagnostic category and across subgroups defined by sex, age and BMI, thereby exploring patterns that may be hidden when considering the sample as a whole.

In our study, individuals with BD exhibited higher levels of *NE* and lower levels of *Extraversion* and *Conscientiousness*. These associations were observed in unadjusted group comparisons and persisted after adjusting for relevant demographic and clinical covariates, including sex, age, and BMI. This pattern closely aligns with a previous meta-analysis of 18 studies comprising 1,694 BD subjects and 2,153 controls, which reported alterations in these three personality domains with concordant effect direction (Hanke et al., 2022). Similarly, a large longitudinal study assessed across two cohorts found consistent results, with elevated *NE* and reduced *Extraversion* and *Conscientiousness* across three time points, whereas *Openness* and *Agreeableness* showed less consistent patterns (Barnett et al., 2011). Importantly, the stability of this personality profile over follow-up assessments suggests that these traits are not merely epiphenomena of acute mood states, but rather reflect relatively stable personality features (Ortelbach et al., 2022; Ryan et al., 2021).

From a dimensional personality perspective, higher *NE* in BD may reflect greater vulnerability to emotional dysregulation, whereas lower *Extraversion* and *Conscientiousness* are consistent with reduced behavioural activation and self-regulatory capacities, respectively (Widiger et al., 2017; Roberts et al., 2014; DeYoung, 2010). From a biological perspective, the personality patterns observed in BD may reflect the convergence of genetic liability, neurobiological changes, environmental and health factors. Genetic studies have suggested overlap between FFM traits and psychiatric disorders, particularly for *NE*, supporting the interpretation of these traits as intermediate phenotypes that may capture part of the biological vulnerability of BD (Grotzinger et al., 2022; Streit et al., 2022; Smeland et al., 2017). Facet-level analyses further mirrored these domain-level findings, with BD participants showing: i) higher scores compared to controls in *NE* related facets, namely *Anxiety*, *Depression*, and *Emotional Volatility*; ii) lower scores in *Extraversion* related facets, namely *Sociability*, *Assertiveness*, and *Energy Level*; and iii) in the *Conscientiousness* domain lower scores in the *Productivity* and *Responsibility* facets, whereas *Organization* did not differ between groups.

While the overall personality profile of BD was characterized by higher *NE* together with lower *Extraversion* and *Conscientiousness*, stratified analyses indicated that *NE* was the most stable component of this profile across sex, age, and BMI subgroups. In contrast, differences in *Extraversion* and *Conscientiousness* appeared more dependent on the subgroup structure.

In sex-stratified analyses, lower *Conscientiousness* in BD was observed only among females, whereas the corresponding effect in males was nominal and did not survive correction for multiple testing. Decreased *Extraversion* was observed in both males and females, although the effect size was larger in the female group. These findings should be interpreted in the context of the limited sex-specific evidence available in the BD personality literature. Most previous Big Five studies in BD have included mixed-sex samples, and sex-stratified analyses remain scarce. One recent study focused exclusively on Han Chinese women with bipolar and unipolar depression (Lin et al., 2025). However, male-only studies have not been performed yet. Thus, our findings extend previous work by suggesting that the reduction in *Conscientiousness* consistently reported in BD is likely not homogenous across sex groups, and more pronounced among females.

Regarding age, BD-related differences in *Extraversion* and *Conscientiousness* did not emerge amongst participants younger than 40 years, but became significantly different in older individuals. This age-dependent pattern should be interpreted cautiously, as previous Big Five studies in BD have rarely examined age-stratified case-control differences. However, it is consistent with broader evidence indicating that Big Five traits vary across adulthood, including age-related changes in *Conscientiousness* and *Openness* (Soto et al., 2011). We also observed an opposite, non-significant trend for *Openness*, with higher scores in younger BD individuals and lower scores in older BD individuals. Although exploratory, this pattern suggests that *Openness* in BD may be shaped by age-related changes in personality expression, generational differences in personality development, or both.

Regarding BMI, BD-related differences in *Extraversion* and *Conscientiousness* were stronger amongst participants with BMI >25 kg/m². In this group, BD individuals showed lower *Extraversion* and *Conscientiousness* than overweight controls, whereas among participants with BMI ≤25 kg/m² these differences were weaker and did not survive correction for multiple testing. This pattern should be interpreted cautiously, particularly for *Conscientiousness*, for which effect sizes were similar across BMI strata but statistical evidence was stronger in the overweight group. Nevertheless, these findings are consistent with previous evidence linking higher BMI and obesity-related outcomes to lower *Conscientiousness* and other personality dimensions in the general population (Bagnjuk et al., 2019; Gerlach et al., 2016; Jokela et al., 2013; Sutin et al., 2011). They are also clinically relevant in BD, where overweight, obesity, and metabolic alterations are highly comorbid (Vancampfort et al., 2015; McIntyre, 2010; McElroy et al., 2002), and may be partially contributing to observed differences between BD cases and controls in *Conscientiousness*. Importantly, recent evidence from the PsyCourse Study further suggests that higher BMI in individuals with BD is associated with poorer cognitive performance, indicating that excess weight may contribute to illness burden not only through obesity-related issues, but also through cognitive and functional impairment (Solé et al., 2025). Together, these findings support the relevance of considering BMI as a health-related modifier of BD-associated personality patterns.

Although *NE* emerged as the most consistent personality alteration in BD, this should be interpreted within a broader transdiagnostic category. Elevated *NE* is not specific to BD, but rather represents a general liability to affective dysregulation that is also commonly observed in other mood and psychiatric disorders (Griffith et al., 2010; Kotov et al., 2010). For instance, comparisons between BD and MDD indicate that both conditions are characterized by increased *NE*, whereas differences in *Extraversion* may contribute to distinguishing bipolar from unipolar depression (Li et al., 2024; Jylhä et al., 2010). Similarly, previous work comparing BD with BPD (Komasi et al., 2022; Renaud et al., 2012) or schizophrenia (Hashimoto et al., 2025) suggests that, although these disorders share broad alterations in personality functioning, they differ in the relative involvement of other domains such as *Extraversion*, *Openness*, and *Agreeableness*.

BD type analyses showed broadly similar BFI-2 profiles between BD-I and BD-II, with the only nominal difference observed for *NE*, which was higher in BD-II. This result is consistent with previous evidence reporting higher *NE* in BD-II than in BD-I (Sparding et al., 2017; Kim et al., 2012). This may reflect the greater depressive burden and affective instability often described in BD-II compared to BD-I (Quilty et al., 2009; Murray et al., 2007).

Several limitations should be acknowledged. First, the cross-sectional design precludes causal inference regarding the directionality of the associations between BD, personality traits, and BMI. Second, although all BD participants were assessed during euthymia, residual mood symptoms may still have influenced self-reported personality scores, particularly *NE*. Third, some subgroup analyses, especially those comparing BD subtypes or stratifying by age and BMI, involved smaller sample sizes and should therefore be interpreted cautiously. Finally, the sample was recruited from a specific clinical and geographical setting, which may limit the generalizability of the findings to more diverse BD populations. Despite these limitations, this study provides an integrated account of personality in BD, emphasizing both the robustness of core trait alterations and the role of clinical and somatic modifiers in shaping their expression. *Conclusions*

In conclusion, this study supports the existence of a robust personality profile associated with BD, characterized by higher *NE* and lower *Extraversion* and *Conscientiousness* compared to controls. These differences were observed both at the domain and facet levels and remained significant after accounting for relevant covariates, suggesting that they may reflect stable dimensions of vulnerability rather than only transient mood-related effects. Importantly, our findings indicate that this personality profile is not homogeneous across all individuals with BD. *NE* emerged as the most consistent alteration across sex, age, and BMI subgroups, whereas differences in *Extraversion* and *Conscientiousness* appeared more dependent on demographic and health factors. The higher *NE* observed in BD-II compared with BD-I further suggests that this domain may capture subtype-sensitive features related to depressive symptomatology. Overall, these results highlight the value of integrating personality assessment into the study of BD heterogeneity. Future longitudinal and multimodal studies will be needed to clarify the causal direction of these associations and their relevance for clinical course, functional outcomes, and personalized interventions.

## CRediT authorship contribution statement

**Lucía Albarracín-García:** Conceptualization, Data curation, Investigation, Methodology, Resources, Validation, Visualization, Writing – review and editing. **Inés García-Ortiz:** Conceptualization, Formal analysis, Investigation, Methodology, Resources, Software, Visualization, Writing – original draft, Writing – review and editing. **Alejandro Porras-Segovia:** Supervision, Writing – original draft, Writing – review and editing. **Laura Navío-García:** Data curation, Resources, Validation, Writing – review and editing. **Laura Jiménez-Muñoz:** Data curation, Resources, Validation, Writing – review and editing. **Elisabet Madridejos-Palomares:** Data curation, Resources, Validation, Writing – review and editing. **Beatriz M. Gonzalez-Toledo:** Data curation, Resources, Validation, Writing – review and editing. **Olatz López-Fernández:** Data curation, Investigation, Supervision, Writing – review and editing. **Enrique Baca-García:** Funding acquisition, Investigation, Project administration, Supervision, Writing – review and editing. **Claudio Toma:** Conceptualization, Funding acquisition, Investigation, Project administration, Resources, Supervision, Writing – review and editing.

## Data Availability

Data are not publicly available. Please contact with the corresponding author for any inquiries

## Acknowledgment

We extend our sincere gratitude to all participants from the MadManic cohort.

