## Supplementary Fig. 1-2 for "Personality Profiles in Bipolar Disorder: Differences Across Diagnostic Subtypes and Associations with Demographic and Health Factors"

### Supplementary Material

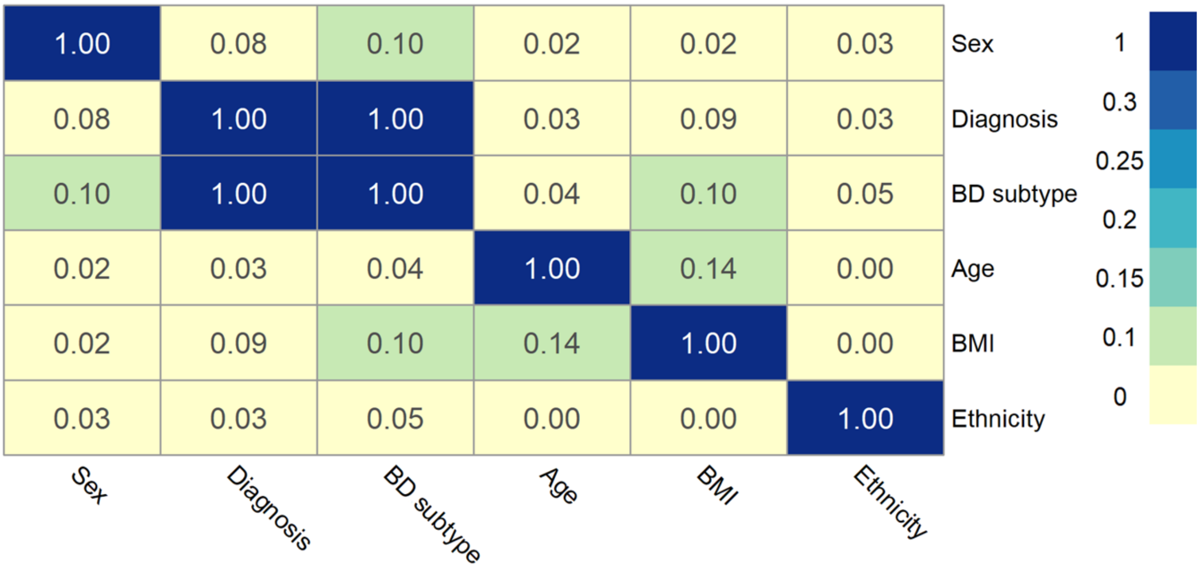


**Supplementary Fig. 1**: Pearson correlation matrix of demographic and clinical covariates across the total sample, including sex, BD diagnosis, BD subtype, age, ethnicity, and BMI).

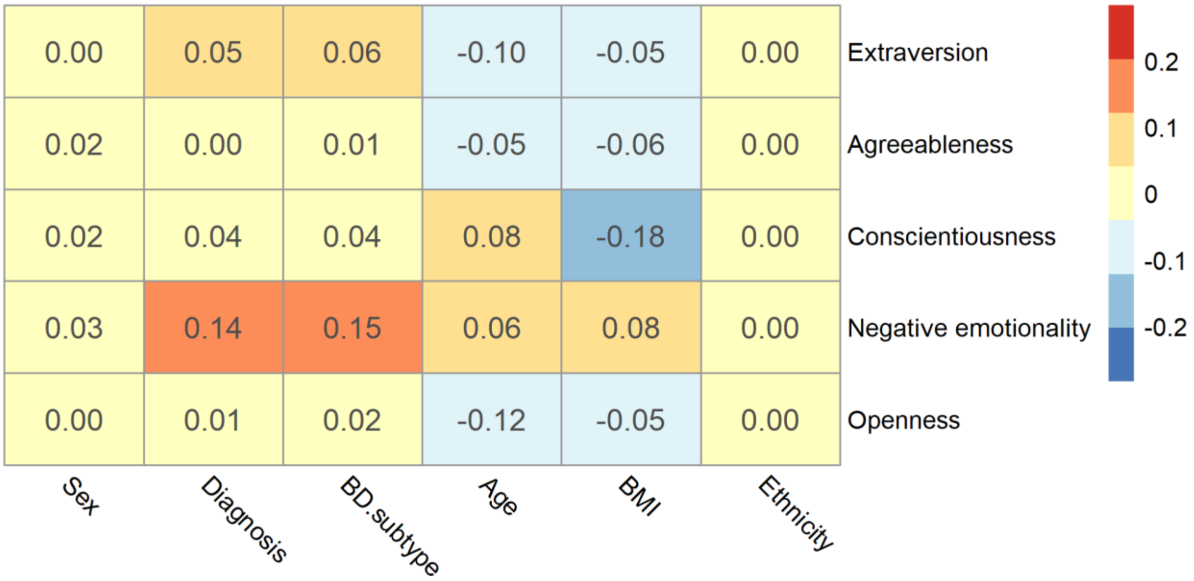


**Supplementary Fig. 2**: Pearson correlation matrix between BFI-2 personality domain scores and demographic and clinical related covariates. Cell values represent Pearson's correlation coefficients.
